# Legionnaires’ disease on the rise: moving from the COVID-19 pandemic towards the 2025 Jubilee in Rome. Data from the regional surveillance system of Lazio Region, Italy (2015-2024)

**DOI:** 10.1101/2025.03.27.25324812

**Authors:** Martina Spaziante, Maurizio D’Amato, Sara Leone, Gabriella De Carli, Gilda Tonziello, Giorgio Nicolò Malatesta, Alessandro Agresta, Claudia De Santis, Valentina Vantaggio, Giovanni Pitti, Maria Concetta Fusco, Pierluca Piselli, Emanuele Nicastri, Claudia Palazzolo, Enrico Girardi, Paola Scognamiglio, Francesco Vairo

**Affiliations:** National Institute for Infectious Diseases “Lazzaro Spallanzani” IRCCS, Via Portuense, 292 - 00149 Rome, Italy

**Keywords:** Infectious diseases, legionellosis, mass gatherings, surveillance

## Abstract

Legionnaires’ disease (LD) is an atypical pneumonia sustained by *Legionella* bacteria. The epidemiological LD scenario in the urban area of Rome (Italy) is unique, moving from facing the COVID-19 pandemic, first capital city in Europe, to the preparation of the 2025 Catholic Jubilee, starting on from December 24, 2024. To provide key determinants for risk assessment in the setting of public health and clinical preparedness activities, we analysed data on LD cases collected from Regional Lazio Surveillance Systems during the 2017-2024 interval period.

## Introduction

Legionnaires’ disease (LD) is an atypical pneumonia sustained by *Legionella* bacteria, transmitted through aerosol inhalation or aspiration of contaminated water (1,2).

Recent years have been characterized by the COVID-19 pandemic, which represented an unprecedented challenge to healthcare systems. Control strategies based on non-pharmaceutical interventions (NPIs) as stay-at-home/lockdown orders and travel restrictions have been extensively implemented (3). Whereas pharmaceutical interventions are known to be disease-specific, NPIs may impact on the transmission of several notifiable infectious diseases (NIDs), not only sustained by air-transmitted pathogens, but also on other NIDs as food-borne diseases, possibly due to behavioral changes secondarily due to stay-at-home policies (4). The impact of NPIs on LD incidence is still under review (5).

In 2024, the city of Rome is preparing for the 2025 Catholic Jubilee, that will take place in Italy starting on 24^th^ December 2024. This event will attract in Rome more than 30 million pilgrims throughout the year (6, 7, 8).

The Jubilee is a mass gathering event which represents a challenge for the regional health system due to both intensity and duration. An increase in notification rates of seasonal NIDs is expected. Additionally, there will be health conditions requiring heightened surveillance and clinical health care assistance during the Jubilee, given the potential rise in risky behaviours. Among these, the excessive overcrowding of hotel facilities could result in compromised hygiene and safety standards, as well as the emergence of makeshift or unauthorized accommodations. Considering this upcoming epidemiological scenario, to provide key determinants of risk assessment in the setting of public health and clinical preparedness activities, we performed an analysis of data on LD cases collected from Regional Lazio Surveillance Systems during the 2017-2024 interval period.

## Methods

A LD confirmed case is defined, according to European Union surveillance definitions published in the *Official Journal of the European Union* (Commission Implementing Decision (EU) 2018/945), as any person with pneumonia with at least one of the following three laboratory criteria:

- isolation of *Legionella spp*. from respiratory secretions or any normally sterile site;
- detection of *Legionella pneumophila* antigen in urine;
- significant rise in specific antibody level to *Legionella pneumophila* serogroup 1 in paired serum samples (9).

LD 2015-2023 data were extracted from LD regional consolidated database. Lazio LD 2024 data were extracted on 7^th^ November 2024 from available sources: 1) national NIDs surveillance system, 2) regional LD Special surveillance system, and, after duplicates cleaning, data underwent record-linkage procedure and consistency checks for cases from both sources. Authors had no access to information that could identify individual participants during or after data collection.

Cases were assigned to week, month and year by symptoms’ onset date. Cases domiciled in or, if data missing, residing in, or if both data missing or if residing outside Lazio Region, notified in Rome City and nearby Fiumicino town were defined “Metropolitan area”, others as “Other”. Current European case definitions were used to classify confirmed or probable cases.

Incidence rate denominator of year i, for i from 2015 to 2024, was calculated as arithmetic mean of Lazio region resident population size as of 1^st^ January 1, of years i and i+1, except for 2024, where i+1 term was numerosity as of 31^st^ August 2024, most recent available data, and resulting arithmetic mean was multiplied by 10/12, because population exposition till 31^st^ October 2024 was assumed. All population data were extracted from National Institute of Statistics demographic data web site (10).

Detection of statistical excess of cases (SEC) in a given week was based on the following method, adapted from (11): a week case frequency is a SEC if the lower bound of its 95% Poisson confidence interval is strictly greater than the arithmetic mean of the previous four-week case frequencies.

Lazio region median weekly rainfall values in millimeters were calculated from weekly distributions of rainfall data recorded by Lazio region weather station network, each weekly distribution obtained summing up the daily rainfall data at weather station level.

Lazio region weekly median temperatures (Celsius scale) were calculated from weekly distribution of median temperatures recorded by Lazio region weather station network, each weekly distribution obtained as median of daily temperature data at weather station level.

Rainfall and temperature daily data by month were downloaded from open data Lazio Region web site (12).

Contingency tables, incidence rates (IR) and incidence rate-ratio (IRR), statistical significance tests, were calculated using R statistical software.

## Results

LD incidence rate (IR) showed an increasing trend from 2015 to 2019, ranging from 2.83 to 4.91 cases per 100,000 person-years (PY). A drop was observed both in 2020 and 2021, followed by a resurgence reaching a peak in 2024, with 390 cases recorded through November 4^th^ 2024 and IR of 8.18 cases per 100,000 PY (Figure 1).

**figure 1.**
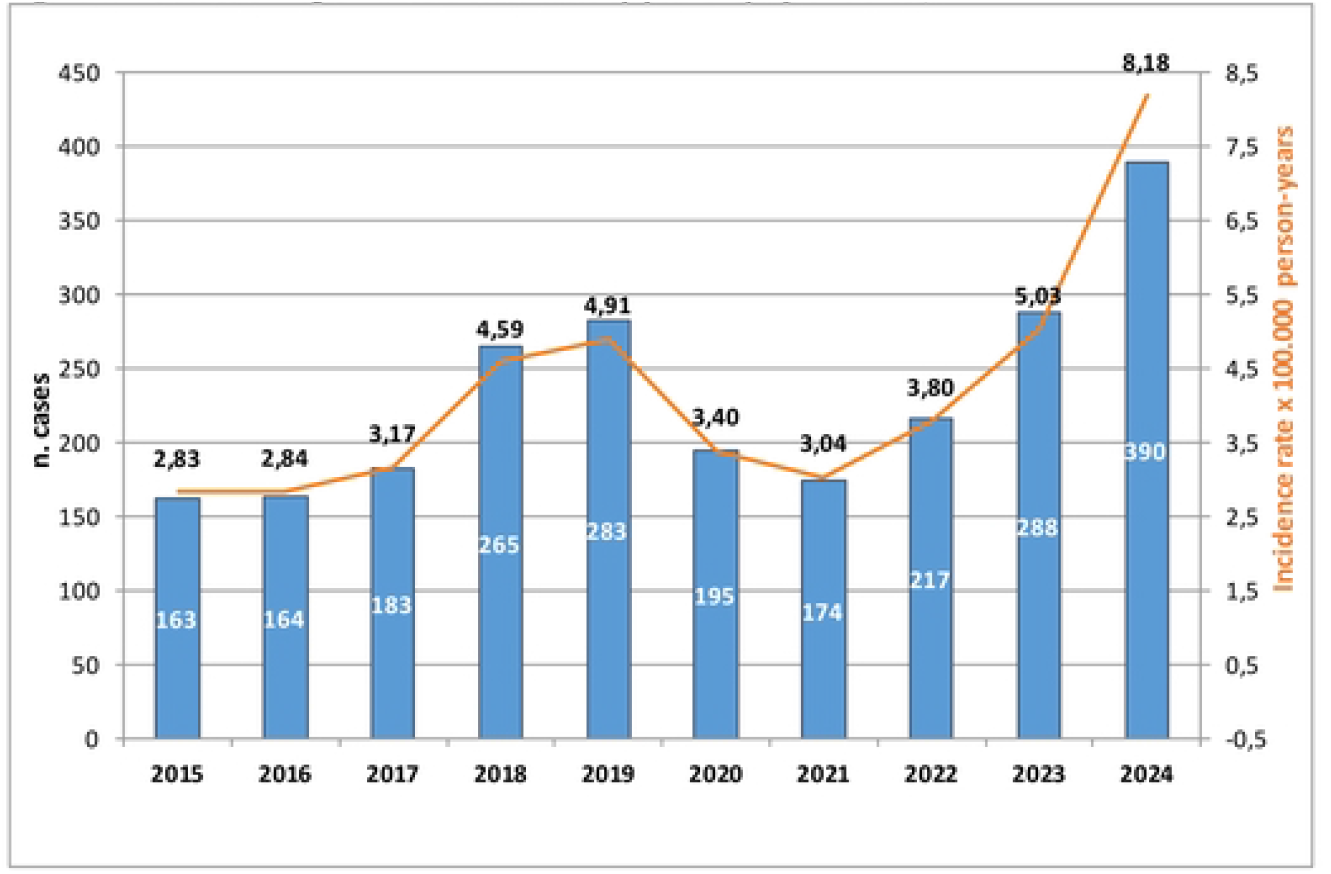
Incidence of legionnaires’ disease cases by year symptoms onset, 2015-2024

Comparison of 2024 LD cases distribution by ISO week to the analogous distributions of 2018, 2019 and 2023, the three years with the highest incidence rate before 2024, showed that, starting from 32^nd^ week, 5^th^-11^th^ August, 2024 largely overcame all the three years, with one week exception. In addition, 2024 was consistently higher in case frequency from 8^th^ week, 19^th^ – 25^th^ February, to 20^th^ week, 13^th^ – 19^th^ May, only tied in 3 weeks. Statistical excesses of cases in 2024 were detected in 28^th^ week, 8^th^ – 14^th^ July, when 14 cases where diagnosed (95% confidence interval (95% CI): 7.7-23.5) versus an average of 4.5 cases in the previous four weeks, and 38^th^ week, 16^th^ – 22^th^ September, when 30 cases occurred (95% CI: 20.2-42.8) versus an average of 12 during the previous four weeks (Figure 2).

**figure 2.**
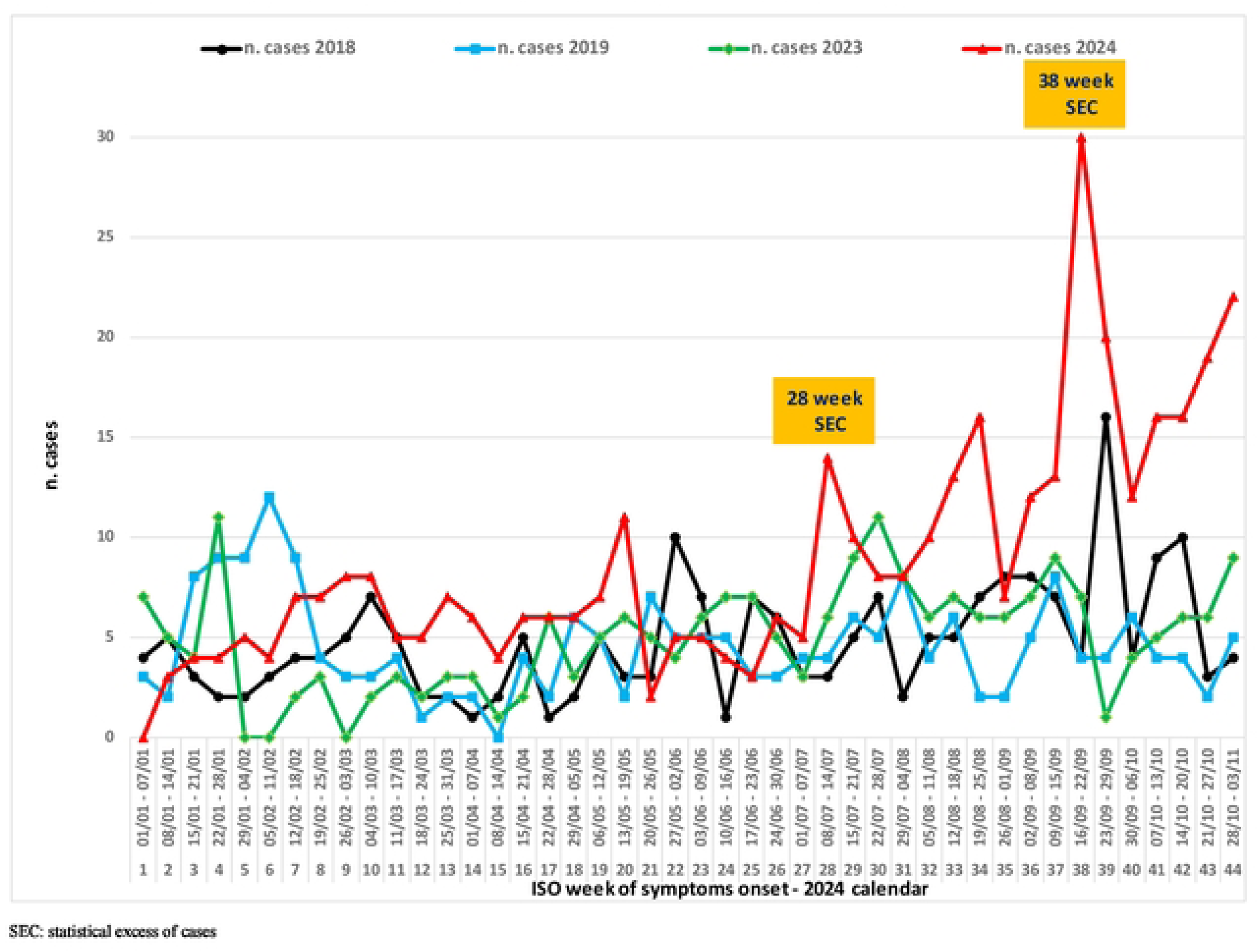
Legionnaries’ disease cases by ISO week of symptoms onset, year 2018, 2019,2023, 2024

An exceptionally high median rainfall week value of 54 mm was registered in 37^th^ which was the maximum value since August 2023, preceded by abundant rainfalls in 36^th^ week too (44 mm, the highest record since April 2024). Between two more modest summer rainfall peaks in the 27^th^ (8 mm) and 33^th^ (11 mm) weeks, a five weeks drought occurred, with only 1 mm of rainfall in the whole period and median maximum temperature consistently above 35°C, peaking at 36.1°C in three weeks. Median maximum temperature declined under 35°C since 34^th^ week, remaining still well above 30°C, and abruptly dropped to 25.6°C in 37^th^ week, reaching 23.5°C in 38^th^ week. Contextually, median minimum temperatures followed a similar pattern, dropping to 13.5°C and 12.9°C in 37^th^ and 38^th^ week, respectively (Figure 3).

**figure 3.**
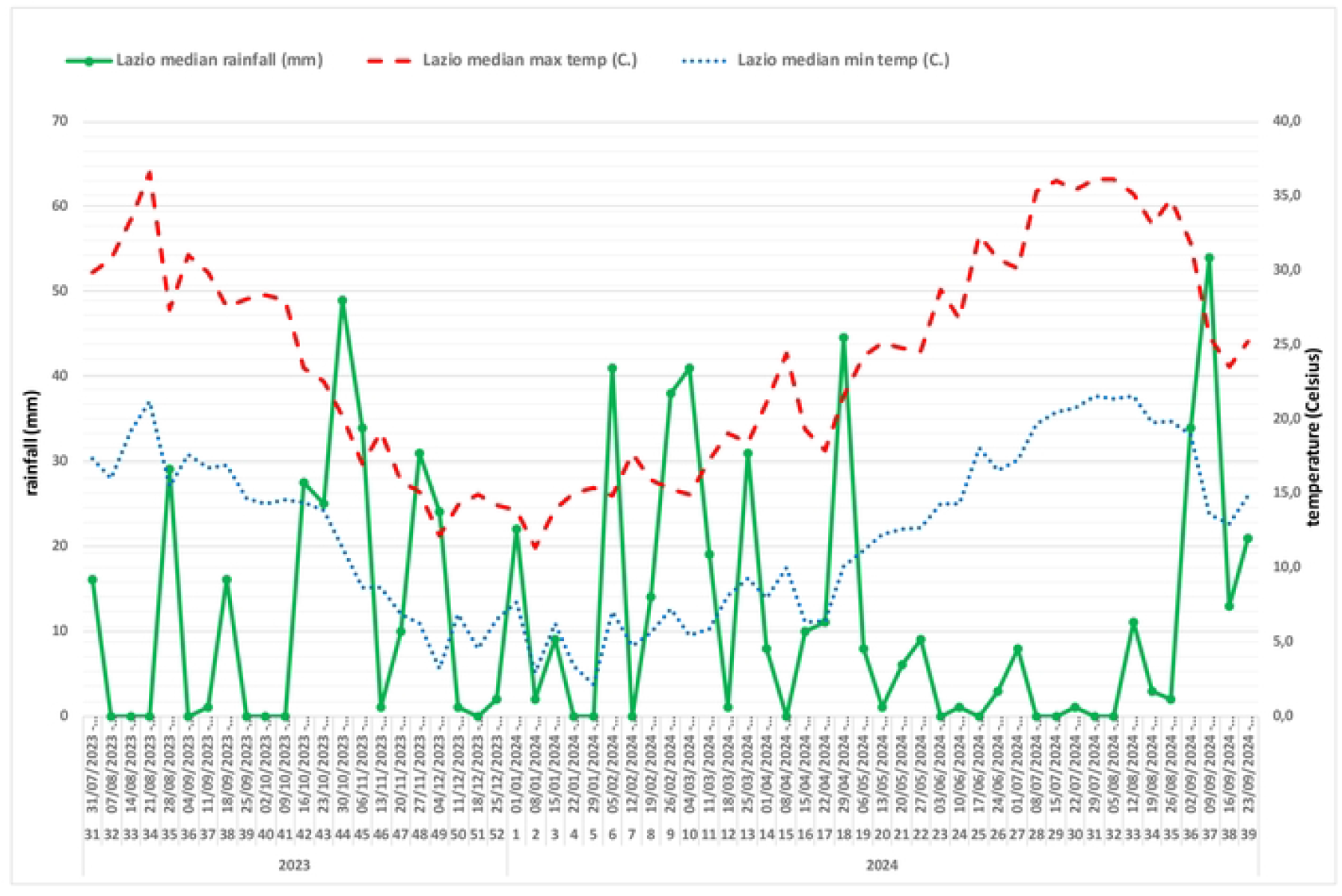
Lazio region median rainfall (millimeter) and median maximum and minimum temperature (Celsius scale) by weeks, Augest 2023-Septemper 2024

During the 2017-2024 interval period, a total of 1,995 confirmed or probable LD cases were reported in the Lazio Region. Overall, the majority of cases occurred in males (67.47%), people aged 50-69 years (43.01%) and living in the metropolitan area (64.06%). The community setting was mostly represented (86.02%). Males accounted for 67.64% of overall LD cases, but this value has swung among years, ranging from the minimum values of 2023 and 2024, 61.46% and 61.79%, respectively, to 74.14% in 2021 (p=0.000). Since 2017 both sexes experienced the lowest and highest incidence in 2021 and 2024, respectively. The occurrence of LD showed different patterns across the age groups during 2017-2024 interval period. While the largest number of annual cases was reported in people aged 50-69 years until 2021, since 2022 the greatest number of cases occurred in those 70 years and older (p=0.001), ranging from 47.00% to 51.03% of cases, in 2022 and 2024, respectively. IR increased with age in each year of 2017-2024 period, rising continuously among adults from 18-49 age class, with IR between 1.10 and 2.65 cases per 100,000 PY, to peak in those 70 years and older particularly in 2024, when an IR of 24.08 cases 100,00 PY was observed in this age class.

LD cases geographical distribution varied significantly across different areas during 2017-2024 period (p=0.006) but metropolitan area consistently registered the highest IR each year, experiencing a peak in 2024, with more than 11 cases per 100,000 PY. Most LD cases usually occurred during the autumn season, and this pattern became more prominent in 2024, with 164 (42.05%) cases recorded during September-November, albeit this quarter weighed around 2/10 months rather 3/12. By assuming 2023 as reference year, 2024 incidence rate ratio (IRR) was 1.63, with the largest increase occurred in people 70 and older (IRR:1.84; 95% CI: 1.47-2.31), in people living in the metropolitan area (IRR: 1.91; 95% CI: 1.57-2.33) or with presumed community exposure (IRR 1.96; 95% CI: 1.66-2.33) (Table 1-2).

**Table 1.** Characteristics of Legionnaires’ disease cases by year of symptoms onset, 2017-2024.

|  | Year of Legionnaires' disease symptoms onset |  |  |  |  |  |  |  |  |  |
| --- | --- | --- | --- | --- | --- | --- | --- | --- | --- | --- |
| Characteristics | Overall, N = 1,995<sup>1</sup> | 2017<br>N = 183<br>(9.2%)<sup>1</sup> | 2018<br>N = 265<br>(13%)<sup>1</sup> | 2019<br>N = 283<br>(14%)<sup>1</sup> | 2020<br>N = 195<br>(9.8%)<sup>1</sup> | 2021<br>N = 174<br>(8.7%)<sup>1</sup> | 2022<br>N = 217<br>(11%)<sup>1</sup> | 2023<br>N = 288<br>(14%)<sup>1</sup> | 2024<br>N = 390<br>(20%)<sup>1</sup> | p-value<sup>2</sup> |
| **Sex** |  |  |  |  |  |  |  |  |  | 0.002 |
| Female | 649 (32.53%) | 50 (27.32%) | 69 (26.04%) | 91 (32.16%) | 70 (35.90%) | 45 (25.86%) | 64 (29.49%) | 111 (38.54%) | 149 (38.21%) |  |
| Male | 1,346 (67.47%) | 133 (72.68%) | 196 (73.96%) | 192 (67.84%) | 125 (64.10%) | 129 (74.14%) | 153 (70.51%) | 177 (61.46%) | 241 (61.79%) |  |
| **Age Median (IQR)** | 66 (55, 78) | 64 (54, 74.50) | 62.00 (52, 77) | 64 (54, 75) | 65 (55, 76) | 68 (56, 77) | 68 (57, 77) | 67 (58, 80) | 70(58, 81) | 0.000 |
| **Age group** |  |  |  |  |  |  |  |  |  | 0.001 |
| 0-17 | 2 (0.10%) | 0 (0.00%) | 1 (0.38%) | 0 (0.00%) | 0 (0.00%) | 0 (0.00%) | 1 (0.46%) | 0 (0.00%) | 0 (0.00%) |  |
| 18-49 | 295 (14.79%) | 32 (17.49%) | 49 (18.49%) | 50 (17.67%) | 34 (17.44%) | 24 (13.79%) | 24 (11.06%) | 35 (12.15%) | 47 (12.05%) |  |
| 50-69 | 858 (43.01%) | 84 (45.90%) | 126 (47.55%) | 125 (44.17%) | 89 (45.64%) | 76 (43.68%) | 90 (41.47%) | 124 (43.06%) | 144 (36.92%) |  |
| 70+ | 840 (42.11%) | 67 (36.61%) | 89 (33.58%) | 108 (38.16%) | 72 (36.92%) | 74 (42.53%) | 102 (47.00%) | 129 (44.79%) | 199 (51.03%) |  |
| **Local Health authority** |  |  |  |  |  |  |  |  |  | 0.006 |
| Metropolitan area | 1,278 (64.06%) | 113 (61.75%) | 182 (68.68%) | 194 (68.55%) | 123 (63.08%) | 106 (60.92%) | 122 (56.22%) | 169 (58.68%) | 269 (68.97%) |  |
| Other | 717 (35.94%) | 70 (38.25%) | 83 (31.32%) | 89 (31.45%) | 72 (36.92%) | 68 (39.08%) | 95 (43.78%) | 119 (41.32%) | 121 (31.03%) |  |
| **Case definition** |  |  |  |  |  |  |  |  |  | 0.000 |
| Confirmed | 1,919 (96.19%) | 168 (91.80%) | 252 (95.09%) | 277 (97.88%) | 191 (97.95%) | 170 (97.70%) | 215 (99.08%) | 267 (92.71%) | 379 (97.18%) |  |
| Probable | 76 (3.81%) | 15 (8.20%) | 13 (4.91%) | 6 (2.12%) | 4 (2.05%) | 4 (2.30%) | 2 (0.92%) | 21 (7.29%) | 11 (2.82%) |  |
| **Setting** |  |  |  |  |  |  |  |  |  | 0.000 |
| Community-acquired | 1,716 (86.02%) | 151 (82.51%) | 224 (84.53%) | 243 (85.87%) | 178 (91.28%) | 147 (84.48%) | 183 (84.33%) | 224 (77.78%) | 366 (93.85%) |  |
| Travel-associated | 139 (6.97%) | 17 (9.29%) | 24 (9.06%) | 19 (6.71%) | 10 (5.13%) | 12 (6.90%) | 18 (8.29%) | 33 (11.46%) | 6 (1.54%) |  |
| Outpatient healthcare-associated | 54 (2.71%) | 4 (2.19%) | 6 (2.26%) | 6 (2.12%) | 3 (1.54%) | 7 (4.02%) | 8 (3.69%) | 17 (5.90%) | 3 (0.77%) |  |
| Hospital-acquired | 86 (4.31%) | 11 (6.01%) | 11 (4.15%) | 15 (5.30%) | 4 (2.05%) | 8 (4.60%) | 8 (3.69%) | 14 (4.86%) | 15 (3.85%) |  |
| **Season** |  |  |  |  |  |  |  |  |  | 0.000 |
| December-February | 381 (19.10%) | 46 (25.14%) | 44 (16.60%) | 73 (25.80%) | 46 (23.59%) | 34 (19.54%) | 45 (20.74%) | 54 (18.75%) | 39 (10.00%) |  |
| March-May | 300 (15.04%) | 27 (14.75%) | 52 (19.62%) | 44 (15.55%) | 18 (9.23%) | 16 (9.20%) | 20 (9.22%) | 43 (14.93%) | 80 (20.51%) |  |
| June- August | 534 (26.77%) | 46 (25.14%) | 63 (23.77%) | 58 (20.49%) | 73 (37.44%) | 52 (29.89%) | 48 (22.12%) | 87 (30.21%) | 107 (27.44%) |  |
| September- November | 780 (39.10%) | 64 (34.97%) | 106 (40.00%) | 108 (38.16%) | 58 (29.74%) | 72 (41.38%) | 104 (47.93%) | 104 (36.11%) | 164 (42.05%) |  |

<sup>1</sup> n (%) except for Age variable<sup>2</sup> Pearson's Chi-squared test; Fisher's Exact test; Kruskal-Wallis rank sum test

**Table 2.** Incidence of Legionnaires’ disease cases by year of symptoms onset, 2017-2024.

|  | Year of Legionnaires' disease symptoms onset |  |  |  |  |  |  |  |  |
| --- | --- | --- | --- | --- | --- | --- | --- | --- | --- |
|  | 2017 | 2018 | 2019 | 2020 | 2021 | 2022 | 2023 | 2024 | 2024 to 2023 |
| Characteristics | IR (95% CI)<sup>1</sup> | IR (95% CI)<sup>1</sup> | IR (95% CI)<sup>1</sup> | IR (95% CI)<sup>1</sup> | IR (95% CI)<sup>1</sup> | IR (95% CI)<sup>1</sup> | IR (95% CI)<sup>1</sup> | IR (95% CI)<sup>1</sup> | IRR (95% CI) |
| **Total population** | 3.17 (2.73-3.66) | 4.59 (4.05-5.18) | 4.91 (4.35-5.52) | 3.40 (2.94-3.91) | 3.04 (2.61-3.53) | 3.80 (3.31-4.34) | 5.03 (4.47-5.65) | 8.18 (7.39-9.04) | 1.63 (1.39-1.90) |
| **Sex** |  |  |  |  |  |  |  |  |  |
| Female | 1.67 (1.24-2.20) | 2.31 (1.80-2.92) | 3.05 (2.46-3.75) | 2.36 (1.84-2.98) | 1.52 (1.11-2.04) | 2.17 (1.67-2.77) | 3.76 (3.10-4.53) | 6.07 (5.13-7.12) | 1.61 (1.25-2.08) |
| Male | 4.78 (4.00-5.66) | 7.03 (6.08-8.09) | 6.90 (5.96-7.95) | 4.51 (3.75-5.37) | 4.66 (3.89-5.54) | 5.52 (4.68-6.47) | 6.39 (5.48-7.40) | 10.43 (9.16-11.83) | 1.63 (1.34-1.99) |
| **Age group** |  |  |  |  |  |  |  |  |  |
| 0-17 |  | 0.11 (0.00-0.6) |  |  |  | 0.11 (0.00-0.62) |  |  |  |
| 18-49 | 1.36 (0.93-1.92) | 2.11 (1.56-2.79) | 2.19 (1.63-2.89) | 1.52 (1.05-2.13) | 1.10 (0.7-1.63) | 1.11 (0.71-1.65) | 1.64 (1.14-2.28) | 2.65 (1.95-3.53) | 1.62 (1.02-2.59) |
| 50-69 | 5.35 (4.26-6.62) | 7.90 (6.58-9.41) | 7.73 (6.43-9.21) | 5.44 (4.37-6.69) | 4.58 (3.61-5.73) | 5.32 (4.28-6.54) | 7.21 (6.00-8.6) | 9.97 (8.41-11.74) | 1.38 (1.08-1.77) |
| 70+ | 7.38 (5.72-9.37) | 9.59 (7.7-11.81) | 11.43 (9.37-13.79) | 7.50 (5.87-9.45) | 7.63 (5.99-9.58) | 10.45 (8.52-12.69) | 13.09 (10.93-15.55) | 24.08 (20.85-27.66) | 1.84 (1.47-2.31) |
| **Local Health authority** |  |  |  |  |  |  |  |  |  |
| Metropolitan area | 3.91 (3.22-4.70) | 6.29 (5.41-7.27) | 6.71 (5.80-7.73) | 4.29 (3.57-5.12) | 3.73 (3.06-4.51) | 4.31 (3.58-5.14) | 5.99 (5.12-6.96) | 11.44 (10.11-12.89) | 1.91 (1.57-2.33) |
| Other | 2.43 (1.89-3.06) | 2.88 (2.29-3.57) | 3.10 (2.49-3.81) | 2.50 (1.96-3.15) | 2.36 (1.83-2.99) | 3.29 (2.66-4.03) | 4.11 (3.40-4.92) | 5.01 (4.16-5.99) | 1.22 (0.94-1.59) |
| **Case definition** |  |  |  |  |  |  |  |  |  |
| Confirmed | 2.91 (2.49-3.38) | 4.36 (3.84-4.94) | 4.81 (4.26-5.41) | 3.33 (2.87-3.83) | 2.97 (2.54-3.45) | 3.76 (3.27-4.30) | 4.67 (4.12-5.26) | 7.95 (7.17-8.79) | 1.70 (1.45-2) |
| Probable | 0.26 (0.15-0.43) | 0.23 (0.12-0.39) | 0.10 (0.04-0.23) | 0.07 (0.02-0.18) | 0.07 (0.02-0.18) | 0.03 (0.00-0.13) | 0.37 (0.23-0.56) | 0.23 (0.12-0.41) | 0.63 (0.27-1.36) |
| **Setting** |  |  |  |  |  |  |  |  |  |
| Community-acquired | 2.62 (2.21-3.07) | 3.88 (3.39-4.42) | 4.22 (3.70-4.78) | 3.10 (2.66-3.59) | 2.57 (2.17-3.02) | 3.20 (2.75-3.7) | 3.92 (3.42-4.46) | 7.68 (6.91-8.51) | 1.96 (1.66-2.33) |
| Travel-associated | 0.29 (0.17-0.47) | 0.42 (0.27-0.62) | 0.33 (0.20-0.51) | 0.17 (0.08-0.32) | 0.21 (0.11-0.37) | 0.31 (0.19-0.50) | 0.58 (0.40-0.81) | 0.13 (0.05-0.27) | 0.22 (0.07-0.53) |
| Outpatient healthcare-associated | 0.07 (0.02-0.18) | 0.10 (0.04-0.23) | 0.10 (0.04-0.23) | 0.05 (0.01-0.15) | 0.12 (0.05-0.25) | 0.14 (0.06-0.28) | 0.30 (0.17-0.48) | 0.06 (0.01-0.18) | 0.21 (0.04-0.73) |
| Hospital-acquired | 0.19 (0.10-0.34) | 0.19 (0.10-0.34) | 0.26 (0.15-0.43) | 0.07 (0.02-0.18) | 0.14 (0.06-0.28) | 0.14 (0.06-0.28) | 0.24 (0.13-0.41) | 0.31 (0.18-0.52) | 1.29 (0.58-2.88) |

<sup>1</sup>cases per 100,000 person-years
IR: incidence rate; IRR: incidence rate ratio; CI: confidence interval

## Discussion

Our analysis highlighted that in the Lazio region, the incidence of LD, which had been steadily increasing since 2015, saw a sharp decline during the first waves of COVID-19 in 2020. However, since 2022, the incidence has gradually risen back to pre-pandemic levels, reaching them in 2023 and surpassing them during the first ten months of 2024. Both pre-pandemic increasing trend and COVID-19-related drop off in LD cases notifications are consistent with literature data (13, 14). This phenomenon may be at least in part explained by the indirect effect of NPIs implementation, including travel restrictions, confirmed by the reduction in travel-associated LD cases registered in 2020 in the Lazio Region.

The pre-pandemic increasing trend was *de facto* restored after the COVID-19 pandemic. To characterize the post-COVID-19 increased notification rates, we observed that in the Lazio region, particularly in 2024, the increase in LD cases notification was driven by sporadic, community-acquired cases, with no clear identification of the environmental source. No significant increase in LD outbreaks was observed in 2024 compared to previous years (16 clusters in 2024 versus 15 in 2023) and local health authorities did not recognize any factor possibly explaining the trend. This is an interesting aspect, as it suggests that the origin of the cases has been difficult to pinpoint, complicating the identification of a single environmental or structural risk factor.

It is likely to recognizes environmental, infrastructural, and population-related drivers. Moffa et al recently conducted a scoping review which demonstrated strong evidence for precipitation as a major driver of LD cases’ increase registered in the USA over the past two decades; temperature, relative humidity, increased testing and improved diagnostic methods were found to be moderate drivers, whereas the ageing population was classified as minor driver (15). The distribution of cases in 2024 shows a significant increase from August to autumn, indicating a typical seasonal pattern linked to climatic conditions that favor the proliferation of *Legionella* bacteria and the risk of exposure (16). Some meteorological factors may have played a role. The impressive 38^th^ week peak in LD cases notification closely followed the drought (week 28^th^-32^th^) and subsequent massive precipitations recorded in weeks 36 ^th^ and 37 ^th^ as well as an abrupt temperature drop since week 37 ^th^. As is known, a period of strong heat followed by intense thunderstorms with a sudden drop in temperature can lead to evaporation phenomena that may have favored the formation of aerosols which, in the presence of sources contaminated by Legionella, may have allowed environmental diffusion with consequent possibility of inhalation (17, 18). It is important to note that within the territory of the municipality of Rome, there are numerous waterways fed by 18 watersheds, the majority of which flow into the Tiber and Aniene rivers that traverse the city (19). Suggestively, the second 2024 statistical excess in LD frequency of cases in week 28^th^ was preceded by a minor rainfall peak in the previous week, and the third summer LD cases’ peak in 34^th^ week coincided with another minor rainfall surge, whereas in both cases no relevant drop in temperature was recorded. The increase in the frequency and intensity of extreme weather events, including droughts followed by torrential rainfall, has been extensively documented in recent decades as a result of global warming. These changes can have a significant impact on ecosystems and public health, particularly with regard to diseases associated with environmental factors, such as Legionnaires’ disease (20).

With regard to demographic variables, according to literature data, also in our epidemiological context, the proportion of Lazio residents ≥ 70 years increased during the observed period, moving from 15.2% in 2015 to 17.3% in 2024 (10). Accordingly, the percentage of elderly LD cases also rose from 42.1% in 2015 to 51.0% in 2024.

## Conclusions

In conclusion, in 2024 the Lazio region experienced an impressive surge of LD cases, driven by sporadic community-acquired cases, affecting particularly the ≥70 yrs old and the metropolitan area of Rome. A clear origin has not yet been identified, whereas meteorological and demographic variables may have played a role. This scenario underscores the need to incorporate climate change considerations into public health policies and mitigation strategies; understanding the complex relationships between environmental, climate, infrastructural and population factors driving LD incidence increase is crucial to optimize mitigation strategies and drive public health policy, particularly in the 2025 Rome Jubilee high-risk scenario. In fact, the insights on LD cases rise just before the inauguration of Jubilee provide to public health authorities a precious risk assessment element stressing the need for timely implementation and strengthening of surveillance, prevention, and infection control activities, as regulated by the relative regional guidelines (21).

## Data Availability

GWAS summary statistics from BBJ and UGR were publicly available (see Table S1). The UK Biobank data was accessed via project 82087 - For access, go to: https://www.ukbiobank.ac.uk/enable-your-research/apply-for-access

## Acknowledgments

The authors would like to thank all patients who contributed data.

